# Omega-3 Supplementation with Lipid Biomarker Assessment in Youth with Migraine: A Quasi-Randomized Controlled Feasibility Trial

**DOI:** 10.64898/2026.09.09.26362637

**Authors:** Jaclyn Bain, Daisy Zamora, Keturah R. Faurot, Saame Raza Shaikh, Anne E. Sanders, Samantha Glover, Klaus Werner, Steven P. Trau, Kimon Divaris, Caroline M. Sawicki

## Abstract

**Background:** Omega-3 polyunsaturated fatty acids may modulate lipid-mediated inflammatory pathways implicated in migraine, yet pediatric trial methodologies evaluating biologic target engagement remain limited. This study evaluated the feasibility of conducting a pediatric nutrition-based migraine intervention trial integrating longitudinal dried blood spot biomarker collection, validated patient-reported outcome measures, dietary assessment, and caregiver-reported acceptability measures.

**Methods:** A quasi-randomized, double-blind, placebo-controlled feasibility trial was conducted at two academic pediatric neurology clinics. Children and adolescents aged 10-17 years with migraine were assigned 1:1 using sequential alternating allocation to daily omega-3 supplementation (850 mg eicosapentaenoic acid + docosahexaenoic acid) or matched coconut oil placebo for 12 weeks. Feasibility outcomes included recruitment yield, participant retention, caregiver-reported supplement adherence, dried blood spot biospecimen completion, 24-hour dietary recall completion, and intervention acceptability assessed via caregiver exit questionnaire. Feasibility benchmarks were established post hoc based on published pediatric trial literature: recruitment >70% of target sample, retention ≥70%, supplement adherence ≥70% of caregivers reporting administration on most days, biospecimen completion ≥90%, dietary recall completion ≥80%, and acceptability rated favorably by ≥70% of caregivers. Clinical efficacy outcomes will be reported separately.

**Results:** Of 58 individuals assessed for eligibility, 57 were enrolled and allocated (29 intervention, 28 placebo). Forty-four participants (77%) completed the 12-week follow-up (22 per group), meeting the retention benchmark. Dried blood spot collection was completed at both time points for all participants who attended study visits (100%). All 44 study completers finished all four planned dietary recalls (100% completion). Caregiver-reported acceptability was high, with the majority rating daily supplement administration as acceptable and reporting openness to future dietary supplement use for migraine management. Thirteen participants discontinued: five withdrew, five were lost to follow-up, two discontinued due to supplement palatability, and one no longer met inclusion criteria. Structured telephone follow-up at weeks 4 and 8 supported retention and adherence monitoring.

**Conclusions:** This feasibility trial demonstrated that a pediatric nutrition-based migraine intervention integrating dried blood spot biomarker collection, repeated dietary recalls, validated patient-reported outcomes, and structured remote follow-up is feasible and acceptable to families. All six post-hoc feasibility benchmarks were met. Findings support progression to a larger-scale randomized controlled trial with modifications including concealed randomization, prospective daily headache diaries, and longer intervention duration.

**Trial registration:** ClinicalTrials.gov, NCT06899074. Registered 20 March 2025.

**Key Messages Regarding Feasibility:** *1) What uncertainties existed regarding the feasibility?:* It was uncertain whether children and adolescents with migraine could be successfully recruited from multiple pediatric neurology clinics, whether families would adhere to daily liquid supplementation over 12 weeks, and whether longitudinal dried blood spot biospecimen collection, repeated telephone-administered dietary recalls, and multidimensional patient-reported outcome assessment could be feasibly integrated within a pediatric outpatient research setting.

*2) What are the key feasibility findings?:* Retention was 77% (44/57), caregiver-reported supplement adherence met the ≥70% threshold, biospecimen and dietary recall completion rates were 100% among study completers, and caregiver-reported acceptability was rated at least moderately acceptable by 96% of study completers. All feasibility benchmarks were met, supporting the overall viability of the study framework. Supplement palatability and disruptions to daily routine were the most commonly reported adherence challenges.

*3) What are the implications of the feasibility findings for the design of the main study?:* A future definitive trial is warranted and should incorporate concealed computer-generated randomization, prospective daily electronic headache diaries as the primary clinical endpoint, longer intervention duration (≥16 weeks), consideration of higher omega-3 doses informed by predictive modeling of omega-3 index response, objective adherence monitoring, expanded lipid biomarker panels, and targeted recruitment strategies to improve demographic diversity.

## Background

Migraine affects an estimated 11% of children and adolescents worldwide and is a leading cause of disability-adjusted life years in youth [1,2]. Pharmacologic prevention remains the mainstay of management, yet network meta-analyses have found limited evidence supporting the efficacy of prophylactic medications in this population [3,4]. These limitations, reflected in the American Academy of Neurology and American Headache Society practice guideline [5], have driven growing clinical and research interest in nutrition-based approaches for pediatric migraine, coinciding with population-level data demonstrating substantial family use of omega-3 fatty acid supplements for children [6,7].

Among dietary strategies, omega-3 polyunsaturated fatty acid (PUFA) supplementation has attracted particular attention. Eicosapentaenoic acid (EPA) and docosahexaenoic acid (DHA) are precursors to specialized pro-resolving mediators and antinociceptive oxylipins involved in trigeminovascular pain signaling, and their blood levels have been shown to correlate with concentrations in the meninges, cranial arteries, and trigeminal ganglia [8–10]. Randomized evidence in adults has shown that increasing omega-3 intake through dietary or supplemental strategies can improve headache outcomes, though the feasibility of delivering such interventions in pediatric populations remains underexplored [11,12]. Harel et al. conducted the only prior pediatric trial of omega-3 supplementation for migraine in 27 adolescents using a crossover design, but the small sample, use of olive oil as a biologically active comparator, and absence of adherence biomarkers limited the ability to draw definitive conclusions [13]. Notably, no prior pediatric study has incorporated objective biomarkers of nutritional target engagement, such as the omega-3 PUFA index, alongside clinical outcome measures, limiting the ability to determine whether supplementation achieves the biologic changes hypothesized to underlie clinical benefit. Recent data demonstrating that blood omega-3 levels correlate with concentrations in trigeminovascular tissues further support the relevance of measuring circulating lipid biomarkers in migraine intervention research [10].

Beyond the limited efficacy data, conducting nutrition-based intervention trials in pediatric populations presents distinct methodological challenges. These challenges include maintaining longitudinal adherence to daily supplementation, integrating repeated dietary assessment and biospecimen collection into study workflows, sustaining participant and caregiver engagement over extended follow-up periods, and coordinating multidimensional outcome assessment within clinical research settings [14–18]. Practical guidance on implementing these procedures in pediatric migraine populations is largely absent from the published literature, and reproducible methodological frameworks are needed to improve the rigor and interpretability of future trials.

Given these uncertainties, a feasibility study was considered a necessary prerequisite to a future, adequately powered randomized controlled trial (RCT). It was unknown whether youth with migraine could be recruited across multiple academic health systems for a nutrition-based intervention, whether families would maintain adherence to daily liquid supplementation over 12 weeks, whether minimally invasive dried blood spot biospecimen collection could be implemented longitudinally in a pediatric outpatient setting, and whether repeated telephone-administered dietary recalls could be feasibly integrated into the study workflow. This study aimed to evaluate the feasibility and acceptability of conducting a quasi-randomized, placebo-controlled trial of omega-3 PUFA supplementation in children and adolescents aged 10-17 years with migraine. Rather than reporting efficacy outcomes, this paper focuses on recruitment, retention, biospecimen and dietary recall completion, supplement adherence, caregiver-reported acceptability, and practical lessons learned during trial implementation. The findings are intended to inform the design of future larger-scale pediatric dietary intervention trials integrating biologic, behavioral, and patient-centered outcome assessment.

## Methods

### Aims and Feasibility Objectives

The primary aim of this study was to evaluate the feasibility and acceptability of conducting a pediatric nutrition-based migraine intervention trial integrating biologic, dietary, clinical, and caregiver-reported outcome measures. The specific feasibility objectives were to evaluate: 1) recruitment of youth with migraine from pediatric neurology clinics across two academic health systems; 2) participant retention over 12 weeks using structured remote follow-up; 3) supplement adherence over 12 weeks assessed via caregiver report during structured telephone follow-up; 4) implementation of longitudinal dried blood spot biospecimen collection in a pediatric outpatient research setting; 5) integration of repeated telephone-administered dietary recalls into the study workflow; and 6) caregiver-reported acceptability of study procedures and daily supplementation. Feasibility objectives were defined prospectively, but formal numerical thresholds were not pre-specified. Progression criteria were therefore established post hoc by benchmarking observed outcomes against published pediatric trial literature, as detailed in the Feasibility Outcomes subsection below. Consistent with guidelines for designing feasibility pilot studies, benchmarks were established to be context-specific and reflective of the demands of the planned full-scale trial [19].

### Study Design and Setting

This was a quasi-randomized, double-blind, placebo-controlled, parallel-group feasibility trial. Children and adolescents aged 10-17 years with migraine were assigned 1:1 using sequential alternating allocation to receive daily omega-3 PUFA supplementation or placebo for 12 weeks. Study assessments were conducted at baseline and at the 12-week follow-up visit, with structured telephone follow-up at weeks 4 and 8.

The study was approved by the Institutional Review Board (IRB) of the University of North Carolina at Chapel Hill (IRB#24-2715, 26 February 2025) and registered at ClinicalTrials.gov (NCT06899074). Recruitment and follow-up procedures were conducted between March 2025 and February 2026. Study reporting followed the Consolidated Standards of Reporting Trials (CONSORT) extension for pilot and feasibility trials [20], adapted for the quasi-randomized design of this study. The completed CONSORT checklist is provided as **Supplemental File 1**.

Participants were recruited from two pediatric neurology clinics affiliated with the University of North Carolina (UNC) Health System and Duke University Health System. A multi-pathway recruitment strategy was used to identify potentially eligible youth while minimizing disruption to routine clinical workflows. The primary identification pathway involved systematic review of upcoming pediatric neurology clinic schedules and electronic medical records through the Carolina Data Warehouse for Health, a centralized institutional data repository. Research personnel screened scheduled patients for preliminary eligibility based on age, documented migraine diagnosis, and absence of exclusionary conditions prior to clinic visits. A secondary pathway involved direct referral from collaborating pediatric neurologists at both institutions, who identified potentially eligible patients during routine clinical encounters. Recruitment materials were also distributed within participating clinics to facilitate self-referral. All study procedures, including informed consent/assent, questionnaire administration, biospecimen collection, and intervention dispensing, were centralized at the UNC Adams School of Dentistry and affiliated clinical research facilities to maintain procedural consistency.

**Figure 1** illustrates the participant workflow and study timeline across the 12-week study period.

**Figure 1.**
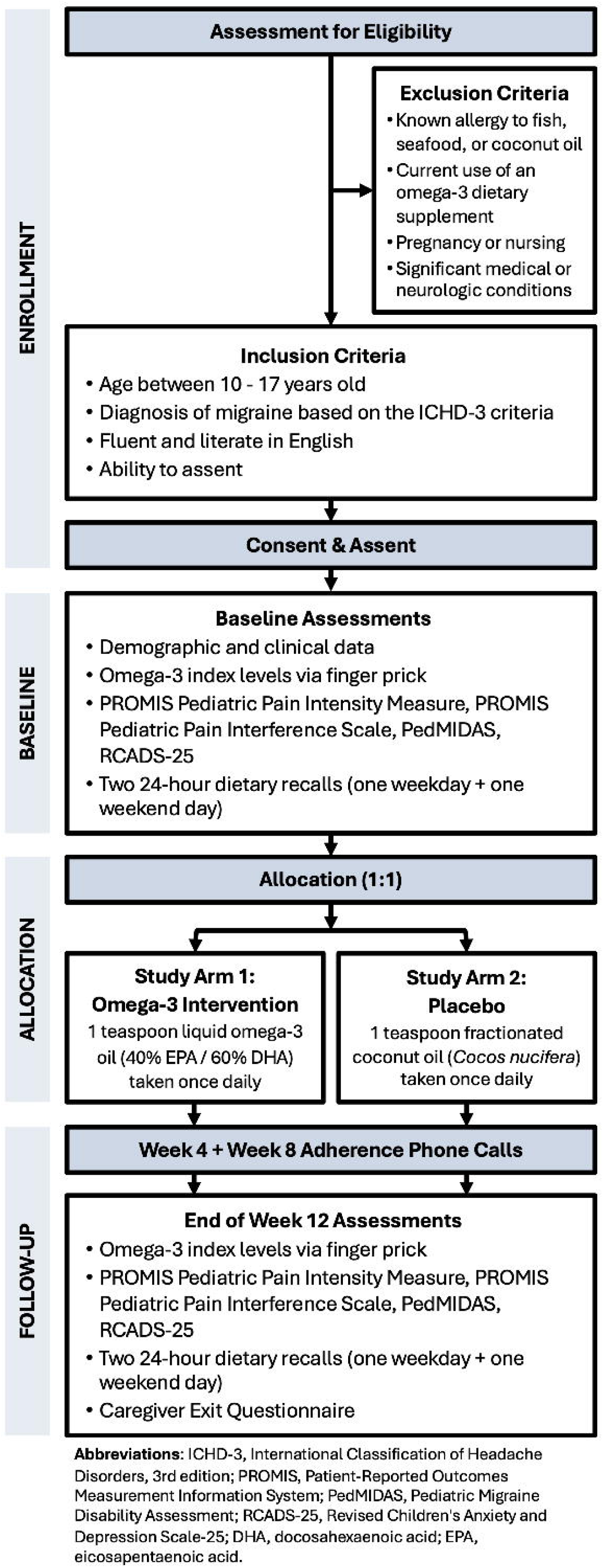
Study design and participant workflow for a 12-week quasi-randomized, double-blind, placebo-controlled feasibility trial of omega-3 polyunsaturated fatty acid supplementation in youth with migraine. Participants were recruited from pediatric neurology clinics, assessed for eligibility, and assigned 1:1 using alternating allocation to daily omega-3 supplementation or coconut oil placebo. Baseline and 12-week assessments included dried blood spot biospecimen collection, validated patient-reported outcome measures, and 24-hour dietary recalls. Structured telephone follow-up was conducted at weeks 4 and 8 to monitor adherence and tolerability. Caregivers completed a caregiver exit questionnaire at the 12-week visit.

### Participants

Families expressing interest completed an initial screening process conducted by study personnel to assess preliminary eligibility. Screening included review of participant age, migraine history and diagnosis, current medication use, relevant medical history, and current omega-3 supplement use. When appropriate, eligibility was further confirmed through review of the electronic medical record and consultation with the participant’s treating neurologist.

Inclusion criteria required participants to be 10-17 years of age with a clinical diagnosis of migraine meeting International Classification of Headache Disorders, 3rd edition (ICHD-3) criteria [21] as confirmed by their current treating pediatric neurologist, English-speaking, and able to complete study procedures with caregiver support. Exclusion criteria included known allergy to fish, seafood, or coconut oil; current use of omega-3 supplementation; significant medical or neurologic conditions that could interfere with study participation or outcome interpretation; and pregnancy or nursing status. The age range of 10-17 years was selected to include children and adolescents capable of providing assent and completing self-report questionnaires while capturing the developmental period during which migraine prevalence increases substantially.

### Allocation and Blinding

Participants were assigned 1:1 to omega-3 PUFA supplementation or placebo using a sequential alternating allocation approach. This method was selected to maintain balanced enrollment across arms throughout the recruitment period, given the small target sample size. Allocation assignments were determined sequentially at the time of enrollment and managed by personnel not involved in outcome assessment. Blinding was maintained through several procedural safeguards. Intervention and placebo formulations were packaged in identical bottles, matched in appearance, and both masked with strawberry flavoring. Study products were dispensed in identical sequentially labeled containers prepared in advance. Group assignment codes were maintained in a separate secure file. Participants, caregivers, investigators, and study personnel involved in data collection and outcome assessment remained blinded to treatment assignment until completion of the primary analyses. Emergency unblinding was permitted only when knowledge of the assigned intervention was essential for clinical management of a serious adverse event or medical emergency. No emergency unblinding was required during the study.

### Intervention and Placebo

Participants in the intervention group received a daily oral omega-3 PUFA supplement (Nordic Naturals, Watsonville, CA, USA). The liquid formulation contained EPA and DHA in a 40:60 ratio, providing 340 mg EPA and 510 mg DHA per teaspoon. A liquid formulation was selected over capsules to facilitate dosing flexibility and improve palatability and swallowability in a pediatric population.

The placebo was a matched daily liquid fractionated coconut oil preparation compounded by Chapel Hill Compounding (Chapel Hill, NC, USA). Fractionated coconut oil, 100% derived from *Cocos nucifera*, was specifically selected because it is composed predominantly of medium-chain and saturated fatty acids that do not participate in omega-3 or omega-6 PUFA metabolic pathways and are not expected to modulate eicosanoid or docosanoid signaling cascades relevant to migraine pathophysiology [22]. This selection addresses a recognized methodological concern in omega-3 trials, as olive oil and corn oil placebos used in prior studies may themselves exert immunomodulatory or metabolically active effects that obscure between-group differences [23].

Participants and caregivers were instructed to administer one teaspoon orally each day, maintain usual dietary habits and migraine management unless medically necessary, and avoid initiating additional omega-3 supplementation. Study products were dispensed immediately following completion of baseline measures with standardized written and verbal administration instructions.

### Data Collection Procedures

#### Lipid biomarker collection

Lipid biomarker collection was performed at baseline and at the 12-week follow-up visit using dried blood spot methodology from OmegaQuant Analytics (Sioux Falls, SD, USA). Finger-prick blood samples were collected by trained study personnel using standardized collection procedures designed to minimize participant discomfort. Following sample collection, dried blood spot cards were air dried and subsequently stored and shipped according to manufacturer and laboratory processing guidelines. Biospecimen analysis was conducted using gas chromatography with flame ionization detection to quantify fatty acid composition. The primary lipid biomarker was the omega-3 PUFA index, defined as the combined percentage of EPA and DHA relative to total identified fatty acids in whole blood. Exploratory lipid biomarkers included the omega-6/omega-3 ratio and the arachidonic acid (AA)/EPA ratio. Dried blood spot methodology was selected to support minimally invasive longitudinal biospecimen collection in pediatric participants while facilitating standardized remote laboratory analysis and simplified specimen handling procedures [24].

#### Dietary Intake Assessment

Dietary intake was assessed at baseline and at the 12-week follow-up using two 24-hour dietary recalls administered for each participant, including one weekday recall and one weekend-day recall at each time point. Dietary recalls were conducted remotely by trained nutrition assessment personnel using the Nutrition Data System for Research (NDSR; Nutrition Coordinating Center, University of Minnesota, Minneapolis, MN, USA). Telephone-administered dietary recalls were used to estimate average intake of total calories, macro- and micronutrients, and dietary fatty acid consumption during the study period. Participants and caregivers were also asked about use of dietary supplements and any substantial changes in dietary habits during the intervention period. The dietary assessment procedures were incorporated to help characterize baseline dietary intake patterns, monitor potential changes in omega-3 fatty acid consumption outside of the study intervention, and evaluate the feasibility of integrating repeated nutrition assessment procedures into pediatric migraine intervention research. Use of both weekday and weekend recalls was intended to improve representation of usual dietary intake patterns across participants.

#### Patient-Reported Outcome Measures

Validated pediatric patient-reported outcome measures were administered at baseline and at the 12-week follow-up visit to assess pain-related symptoms, migraine-related disability, and psychological distress. Questionnaires were completed electronically using standardized study data collection procedures.

Pain intensity and pain interference were assessed using the Patient-Reported Outcomes Measurement Information System (PROMIS) Pediatric Pain Intensity [25] and PROMIS Pediatric Pain Interference [26] short forms. These instruments are validated for use in pediatric populations and generate standardized T-scores, with higher scores reflecting greater symptom severity or functional impact.

Migraine-related disability was assessed using the Pediatric Migraine Disability Assessment (PedMIDAS), a validated questionnaire evaluating migraine-related impairment in school, home, and social functioning over the preceding three months [27]. Psychological distress symptoms were assessed using the Revised Children’s Anxiety and Depression Scale short version (RCADS-25), a validated pediatric measure of anxiety- and depression-related symptoms [28]. Pediatric health-related quality of life was planned to be assessed using the validated Pediatric Quality of Life Inventory (PedsQL) Measurement Model [29].

Patient-reported outcome measures were selected to support multidimensional assessment of migraine-related symptom burden and psychosocial functioning while evaluating the feasibility of integrating standardized pediatric outcome instruments into nutrition-based migraine intervention research.

#### Caregiver Exit Questionnaire

At the 12-week follow-up visit, caregivers completed a study-specific caregiver exit questionnaire designed to assess intervention acceptability, perceived feasibility of study participation, and caregiver perspectives regarding non-pharmacologic migraine management approaches. Questionnaire items evaluated caregiver perceptions of daily supplement administration, openness to future dietary supplement use for migraine management, perceived changes in migraine symptoms during the study period, and overall experience with study participation.

Additional questionnaire items assessed caregiver perspectives regarding access to non-pharmacologic migraine treatment approaches, concerns related to currently available migraine management options, and perceived barriers to implementation of nutrition-based interventions in pediatric migraine care.

The caregiver exit questionnaire was incorporated to evaluate caregiver-centered feasibility and acceptability outcomes relevant to future pediatric dietary intervention trials. Inclusion of caregiver-reported measures was considered particularly important given the role of caregivers in supplement administration, longitudinal participation, and treatment decision-making in pediatric populations.

#### Telephone Follow-Up and Adherence Monitoring

Structured telephone follow-up procedures were conducted by trained study personnel at weeks 4 and 8 following allocation. Follow-up calls were designed to support participant retention, monitor intervention feasibility, and identify any concerns related to supplement administration or study participation during the intervention period.

During each follow-up call, caregivers were asked standardized questions regarding daily supplement administration, missed doses, tolerability, and challenges related to adherence. Study personnel also collected information regarding interval changes in medical history, migraine management, medication use, and adverse events. Caregivers were additionally provided opportunities to ask questions regarding study procedures or supplement administration.

Telephone follow-up procedures were incorporated to support longitudinal engagement and remote monitoring while minimizing participant burden associated with additional in-person study visits. These procedures also allowed ongoing assessment of the feasibility of integrating caregiver-supported adherence monitoring within a pediatric nutrition-based intervention framework. No formal pill count or bottle weight measurement was performed; adherence assessment relied on caregiver report, consistent with the feasibility design and the goal of minimizing participant burden.

### Feasibility Outcomes

Feasibility benchmarks were informed by empirical data from comparable pediatric trials, methodological frameworks for nutrition RCTs, and clinical reasoning specific to the study population and intervention [14,16,19,30,31]. Feasibility domains and assessment methods were defined prospectively as part of the study protocol. Numerical benchmarks for each domain were established post hoc by reference to published pediatric trial literature, as formal progression criteria were not pre-specified prior to enrollment. Feasibility was evaluated across six domains:

- **Recruitment:** Ability to enroll the target sample from pediatric neurology clinics across two academic health systems within the planned recruitment period. Benchmark: enrollment of ≥70% of the target sample (n=80). This threshold is context-specific and reflects the demands of the planned full-scale trial, as recommended by Teresi et al [19], as well as considering well-documented recruitment challenges in pediatric clinical trial including parental decision-making, socioeconomic factors, and study burden identified as key barriers [32].
- **Retention:** Proportion of enrolled participants completing the 12-week follow-up. Benchmark: ≥70% retention, informed by published data indicating median retention of 92% in pediatric RCTs, with higher retention observed in trials of shorter duration and those using active or placebo comparators [30]. The lower threshold accounts for the additional participant burden associated with biospecimen collection and repeated dietary recalls in a pediatric population where time constraints, transportation barriers, and complexity of consent/assent procedures are well-documented contributors to attrition in nutrition and migraine trials [15,17,18,33].
- **Biospecimen Completion:** Proportion of attending participants with paired baseline and 12-week dried blood spot samples of sufficient quality for laboratory analysis. Benchmark: ≥90% completion among participants attending both visits, consistent with evidence supporting the feasibility of dried blood spot methodology in pediatric and community-based research settings [34,35] as well as erythrocyte or whole blood EPA/DHA measurement being essential for objective verification of omega-3 adherence [24,36].
- **Dietary Recall Completion:** Proportion of participants completing all four planned 24-hour dietary recalls. Benchmark: ≥80% completion among study completers, acknowledging recognized scheduling challenges associated with telephone-administered dietary recalls in pediatric populations, particularly for weekend-day recalls [17,18]. This benchmark assists in determining the feasibility and acceptability of the dietary assessment procedure in this pediatric population, to characterize the distribution of background dietary omega-3 intake for informing eligibility criteria and stratification in the full-scale trial, and to evaluate concordance between caregiver-reported dietary intake and the objective blood-based omega-3 biomarker, a comparison that the International Society for the Study of Fatty Acids and Lipids (ISSFAL) recommends as essential in all omega-3 supplementation research [24,37].
- **Adherence:** Caregiver-reported daily supplement administration during structured telephone follow-up at weeks 4 and 8. Benchmark: >70% of caregivers reporting administration of the supplement on most days (>5 of 7 days per week), informed by systematic review evidence indicating that adherence to daily oral supplementation in pediatric populations typically ranges from 70% to 90% when assessed by caregiver report, with lower rates observed in trials of longer duration and those involving supplements with palatability challenges [15,38]. This benchmark was selected to reflect a clinically meaningful level of supplement exposure while accounting for the known challenges of daily liquid fish oil administration in children, including taste, odor, and disruptions to routine.
- **Acceptability:** Caregiver-reported acceptability of daily supplementation and study procedures on the caregiver exit questionnaire. Benchmark: ≥70% of caregivers rating study participation and supplement administration as acceptable, informed by growing clinical and research interest in non-pharmacologic and nutraceutical approaches for pediatric migraine given the limited evidence base supporting current pharmacologic options [39] and the central role of caregivers in supplement administration and treatment decision-making in pediatric populations [15]. This benchmark also accounts for the known palatability challenges of fish oil supplementation in children (taste, odor, capsule size), informed by the systematic review evidence showing that while the majority of pediatric nutrition interventions achieve high acceptability, a substantial proportion of caregivers report concerns related to gastrointestinal side effects and taste [15,18].

### Safety Monitoring

During each study contact (weeks 4, 8, and 12), caregivers were asked standardized questions regarding any adverse events, new medical diagnoses, emergency department visits, hospitalizations, or changes in medication use since the last contact. Adverse events were documented in the secure study database and classified by severity, relatedness to the study intervention, and expectedness. Serious adverse events were to be reported to the UNC IRB in accordance with institutional reporting requirements.

Given the feasibility nature of the study, the low-risk profile of omega-3 supplementation at the administered dose, and the small sample size, a formal Data Safety Monitoring Board was not convened. Safety oversight was maintained by the principal investigator and study team through regular review of enrollment progress, retention, adverse event reports, and protocol deviations. Known risks associated with omega-3 supplementation were reviewed during the consent process and included the potential for allergic reactions in individuals with fish allergies and mild gastrointestinal symptoms such as nausea, abdominal discomfort, or fishy aftertaste. To minimize risk, individuals with a known fish or seafood allergy were excluded from participation.

Participants were permitted to discontinue the study intervention at any time for any reason, including caregiver or participant preference, intolerance, or adverse effects. No dose modifications were planned, as the intervention consisted of a fixed daily dose. Discontinuation of the intervention by the study team was permitted if a participant experienced a clinically significant adverse event judged to be related to the study product, developed a new medical condition meeting exclusion criteria (e.g., pregnancy), or if continued participation was deemed unsafe by the principal investigator in consultation with the participant’s treating neurologist.

### Sample Size

The target enrollment was 80 participants (40 per group). This sample size provides sufficient data to evaluate the feasibility objectives described above, including recruitment yield across two academic health systems, retention over 12 weeks, completion rates for biospecimen collection and dietary recalls, caregiver-reported adherence, and intervention acceptability, while generating adequate variability in feasibility metrics to inform the design of a future full-scale trial. The target of 40 per group meets or exceeds published recommendations for pilot and feasibility trials, which suggest a minimum of 30-36 participants per arm to estimate key design parameters with reasonable precision [40,41]. The target of 40 per group also accounted for anticipated attrition of approximately 20-30% over the 12-week participation period, ensuring that a minimum of 21 completers per group would be available for the companion biomarker analyses. With a two-sided significance level of 0.05 and a standardized effect size of 0.865, informed by predictive modeling of omega-3 index response to supplementation, 21 participants per group was estimated to provide 80% power to detect a statistically significant change in the omega-3 PUFA index from baseline to post-intervention. Consistent with the feasibility design, biomarker comparisons are reported descriptively and should be interpreted as preliminary estimates to inform the definitive trial rather than as confirmatory efficacy findings. The results of the assessment of biologic target engagement are reported in a separate companion manuscript.

### Statistical Analysis

Descriptive statistics were used to summarize demographic characteristics, feasibility metrics, intervention acceptability measures, and study completion rates. Continuous variables were summarized using means and standard deviations or medians and interquartile ranges, as appropriate, while categorical variables were summarized using frequencies and percentages. Feasibility outcomes were evaluated against the post-hoc benchmarks described above. Questionnaire responses from the caregiver exit questionnaire were summarized descriptively to evaluate caregiver-reported feasibility and acceptability of study procedures and intervention participation.

Clinical and biologic outcome data, including changes in the omega-3 PUFA index and patient-reported outcome measures, are reported in a separate companion manuscript. In that manuscript, changes from baseline to the 12-week follow-up were evaluated using analysis of covariance (ANCOVA) models adjusting for baseline values, with adjusted mean differences and 95% confidence intervals estimated for between-group comparisons. These analyses were exploratory and hypothesis-generating in nature; the study was not powered to detect clinically meaningful between-group differences in clinical outcomes, and any such results should be interpreted with caution. Statistical analyses were performed using Stata version 19 SE, and statistical significance was evaluated using two-sided tests with an alpha level of 0.05.

### Data Management and Confidentiality

Study data were collected and managed using the Carolina Data Acquisition and Reporting Tool (CDART), a secure, Health Insurance Portability and Accountability Act (HIPAA) compliant electronic clinical research database maintained by the University of North Carolina at Chapel Hill. CDART meets federal Part 11 guidelines for clinical study data acquisition and was used to collect participant screening information, demographic data, medical history, questionnaire responses, and study tracking information.

Participants were assigned unique study identification numbers to support de-identification of research data and biospecimens. Identifiable participant information and linkage files were stored separately from study outcome data on secure institutional servers with access restricted to authorized IRB-approved study personnel who had completed required HIPAA and human subjects research training. All study procedures involving participant confidentiality, data security, and research record management were conducted in accordance with institutional policies, applicable federal regulations, and approved institutional review board protocols governing research involving pediatric participants.

## Results

### Recruitment and Enrollment

Of 58 individuals assessed for eligibility, 57 were enrolled and allocated (71% of the target sample of 80), with one participant excluded due to medical history criteria. The multi-pathway recruitment strategy yielded participants from both the Carolina Data Warehouse screening pathway and direct clinician referral, though the relative yield of each pathway varied. Data warehouse screening enabled systematic identification of potentially eligible patients in advance of scheduled neurology clinic visits and was the primary source of recruitment volume. Direct clinician referral from collaborating pediatric neurologists provided a complementary pathway that identified patients who may not have been captured through electronic record screening alone. Self-referral through clinic-based recruitment materials generated limited additional enrollment.

Coordination across two academic health systems (UNC and Duke) required ongoing communication between research personnel and clinical teams at each site. Centralizing all study procedures at UNC facilitated procedural consistency but required participants recruited through Duke to travel to UNC for in-person visits, which may have limited enrollment from that site. The sample size was smaller than the original target of 40 per group due to a fixed, prespecified enrollment window. While this limits the precision of effect estimates, the achieved sample met the minimum threshold necessary to address the feasibility objectives of this trial. The recruitment benchmark (≥70% of target sample) was met.

**Figure 2** presents the CONSORT flow diagram for participant screening, enrollment, allocation, and follow-up

**Figure 2.**
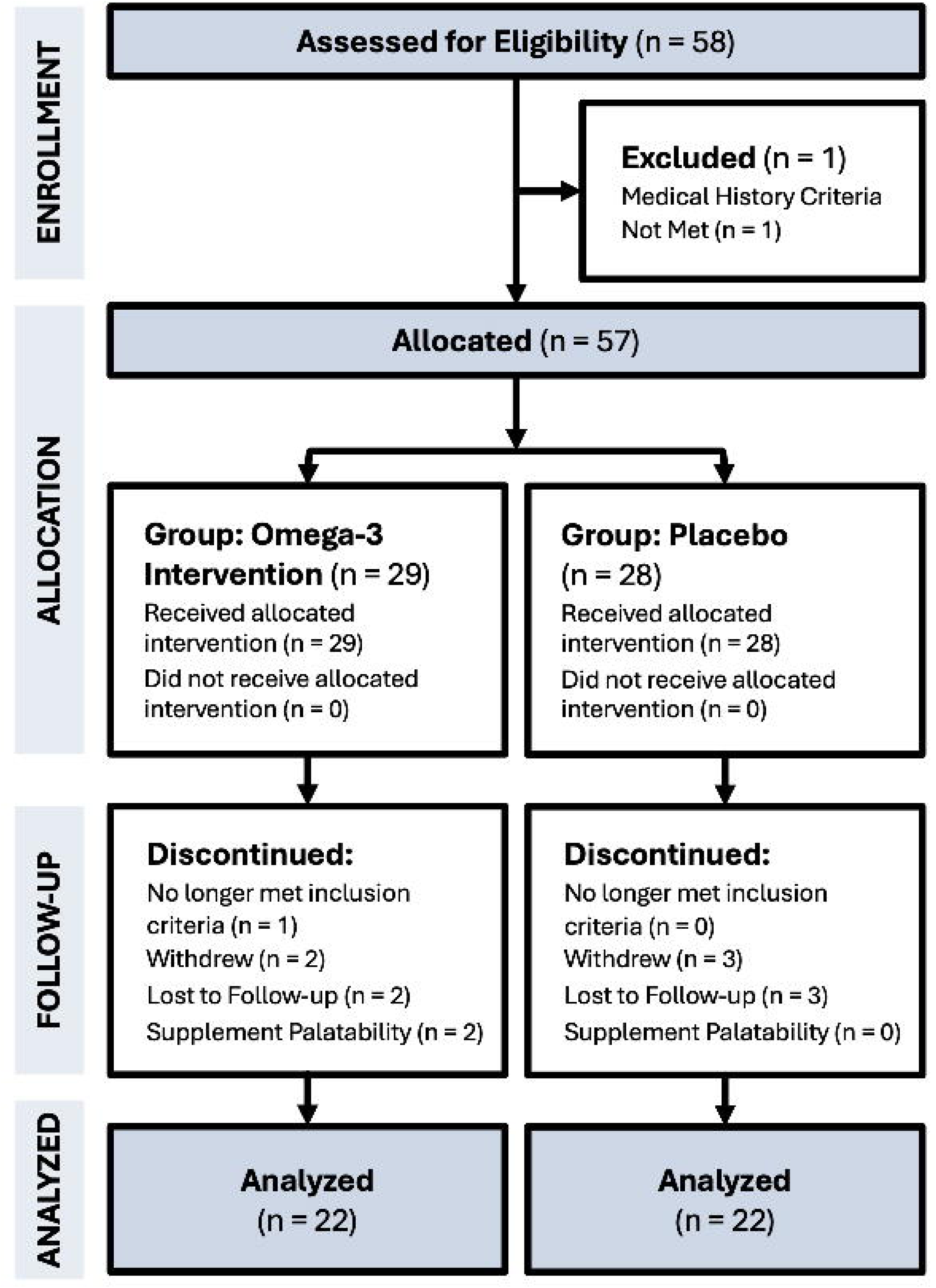
CONSORT flow diagram of participant enrollment, allocation, follow-up, and analysis. A total of 58 individuals were assessed for eligibility, of whom 57 were allocated to the omega-3 PUFA intervention group (n=29) or placebo group (n=28). Thirteen participants discontinued during the 12-week intervention period. A total of 44 participants (22 per group) completed the 12-week follow-up and were included in the feasibility analysis.

### Retention and Reasons for Discontinuation

Of 57 enrolled participants, 44 (77%) completed the 12-week follow-up visit and were included in the final analysis (22 per group). Thirteen participants discontinued after initiating the intervention: five withdrew from study participation, five were lost to follow-up, two discontinued due to difficulties related to supplement palatability or taste, and one no longer met inclusion criteria. Participants who completed the study were similar in age and sex distribution to those who discontinued. Among the 13 non-completers, 69% were female, 15% were male, 15% did not disclose gender, the mean age was 14.8 years, and 8% were recruited via the Duke University Health System pathway. Baseline demographic characteristics of study completers are summarized in **Table 2**. Both palatability-related discontinuations occurred in the active intervention group. No clear demographic or clinical pattern distinguished completers from non-completers, though the small number of discontinuations limits interpretation. Attrition was somewhat higher in the intervention group (7 of 29, 24%) compared with the placebo group (6 of 28, 21%), though the study was not designed to formally compare discontinuation rates. The structured telephone follow-up calls at weeks 4 and 8 appeared to support retention by maintaining regular contact with families and providing opportunities to address concerns early in the study period. Flexible scheduling for the 12-week visit also facilitated visit completion. The retention benchmark (≥70%) was met.

**Table 1.**
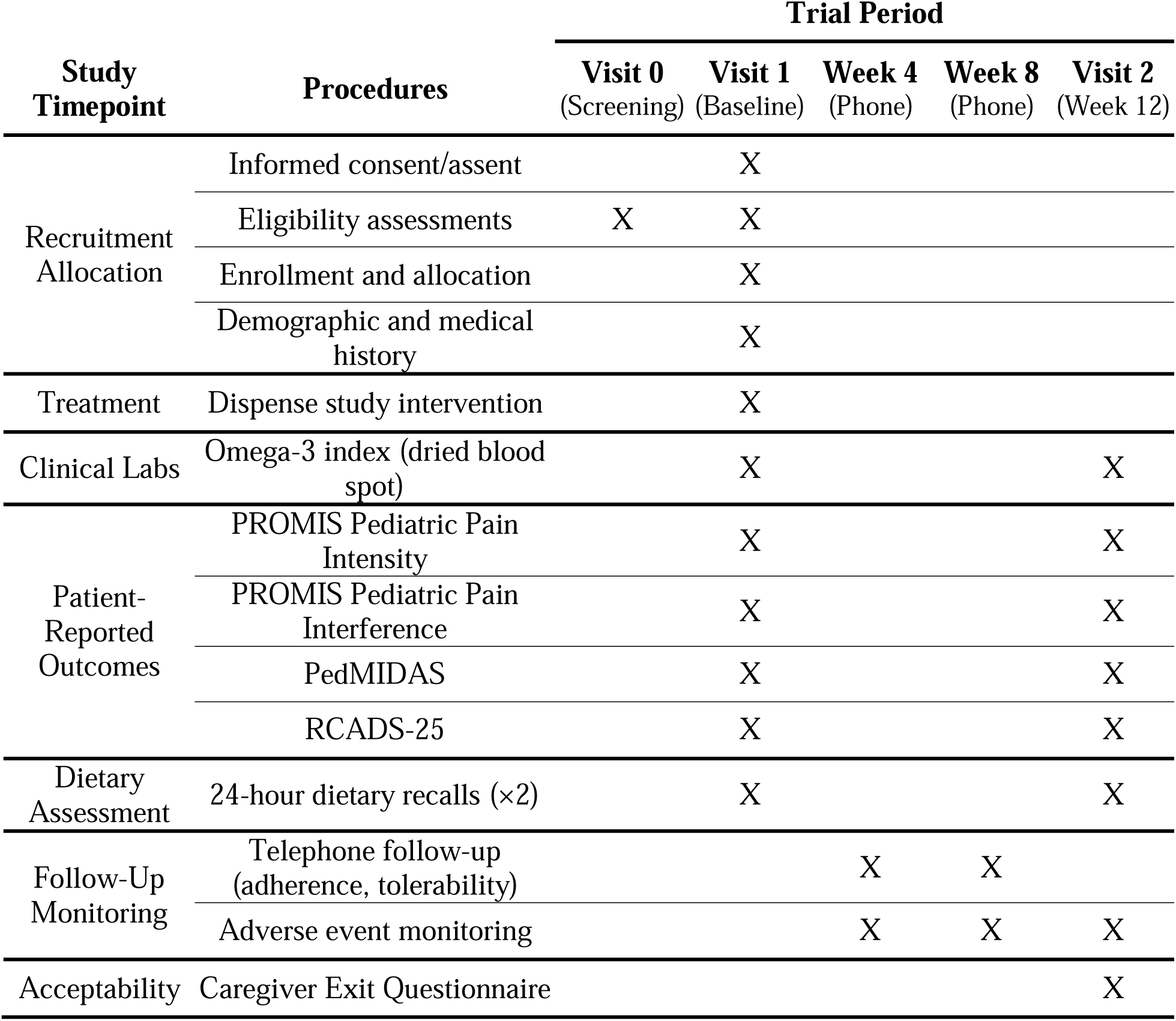
summarizes the schedule of study procedures and assessments across the trial period.

| Study Timepoint | Procedures | Trial Period |  |  |  |  |
| --- | --- | --- | --- | --- | --- | --- |
|  |  | Visit 0<br>(Screening) | Visit 1<br>(Baseline) | Week 4<br>(Phone) | Week 8<br>(Phone) | Visit 2<br>(Week 12) |
| Recruitment<br>Allocation | Informed consent/assent |  | X |  |  |  |
|  | Eligibility assessments | X | X |  |  |  |
|  | Enrollment and allocation |  | X |  |  |  |
|  | Demographic and medical history |  | X |  |  |  |
| Treatment | Dispense study intervention |  | X |  |  |  |
| Clinical Labs | Omega-3 index (dried blood spot) |  | X |  |  | X |
| Patient-Reported<br>Outcomes | PROMIS Pediatric Pain Intensity |  | X |  |  | X |
|  | PROMIS Pediatric Pain Interference |  | X |  |  | X |
|  | PedMIDAS |  | X |  |  | X |
|  | RCADS-25 |  | X |  |  | X |
| Dietary<br>Assessment | 24-hour dietary recalls (×2) |  | X |  |  | X |
| Follow-Up<br>Monitoring | Telephone follow-up (adherence, tolerability) |  |  | X | X |  |
|  | Adverse event monitoring |  |  | X | X | X |
| Acceptability | Caregiver Exit Questionnaire |  |  |  |  | X |

**Table 2.**
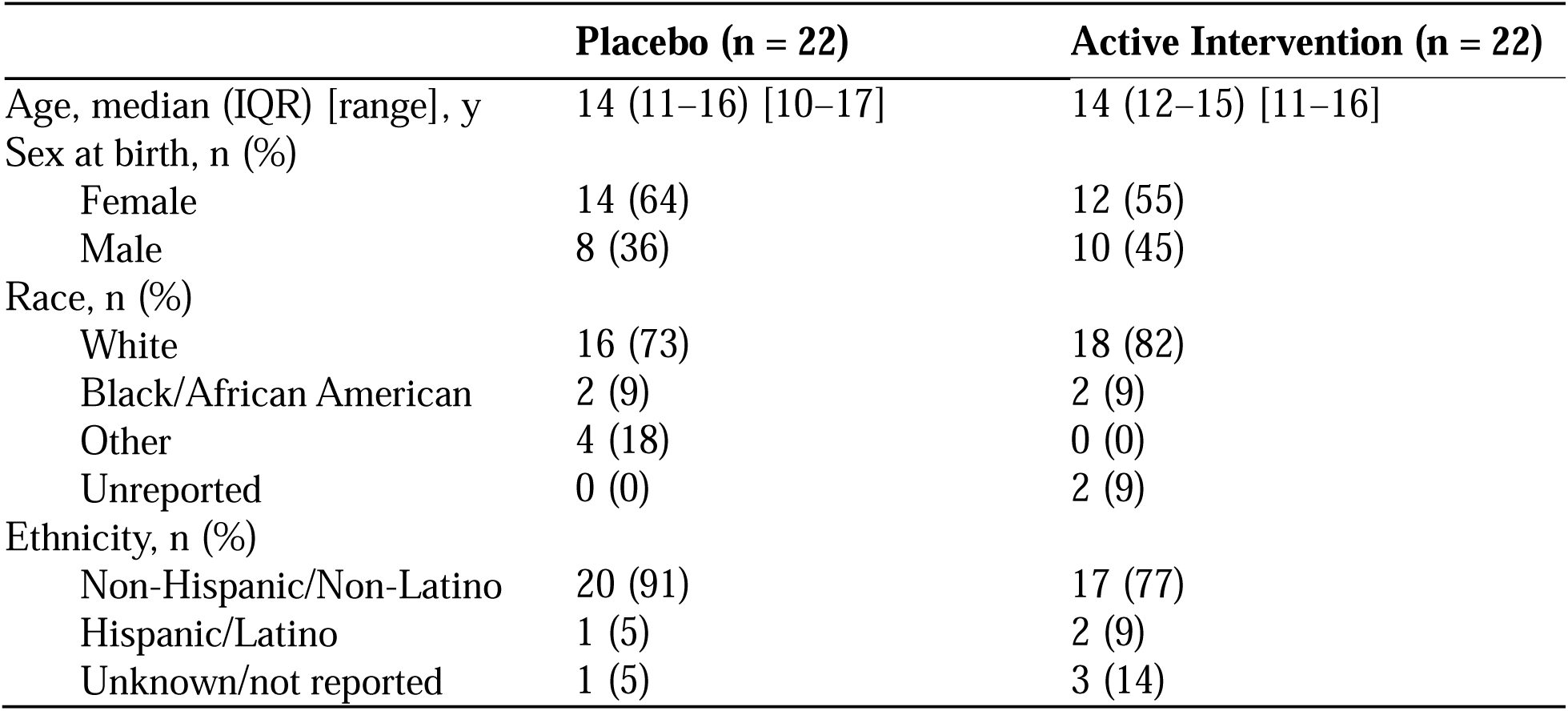
Comparison of demographic characteristics at baseline in the two treatment groups.

|  | Placebo (n = 22) | Active Intervention (n = 22) |
| --- | --- | --- |
| Age, median (IQR) [range], y | 14 (11–16) [10–17] | 14 (12–15) [11–16] |
| Sex at birth, n (%) |  |  |
| Female | 14 (64) | 12 (55) |
| Male | 8 (36) | 10 (45) |
| Race, n (%) |  |  |
| White | 16 (73) | 18 (82) |
| Black/African American | 2 (9) | 2 (9) |
| Other | 4 (18) | 0 (0) |
| Unreported | 0 (0) | 2 (9) |
| Ethnicity, n (%) |  |  |
| Non-Hispanic/Non-Latino | 20 (91) | 17 (77) |
| Hispanic/Latino | 1 (5) | 2 (9) |
| Unknown/not reported | 1 (5) | 3 (14) |

**Table 3.**
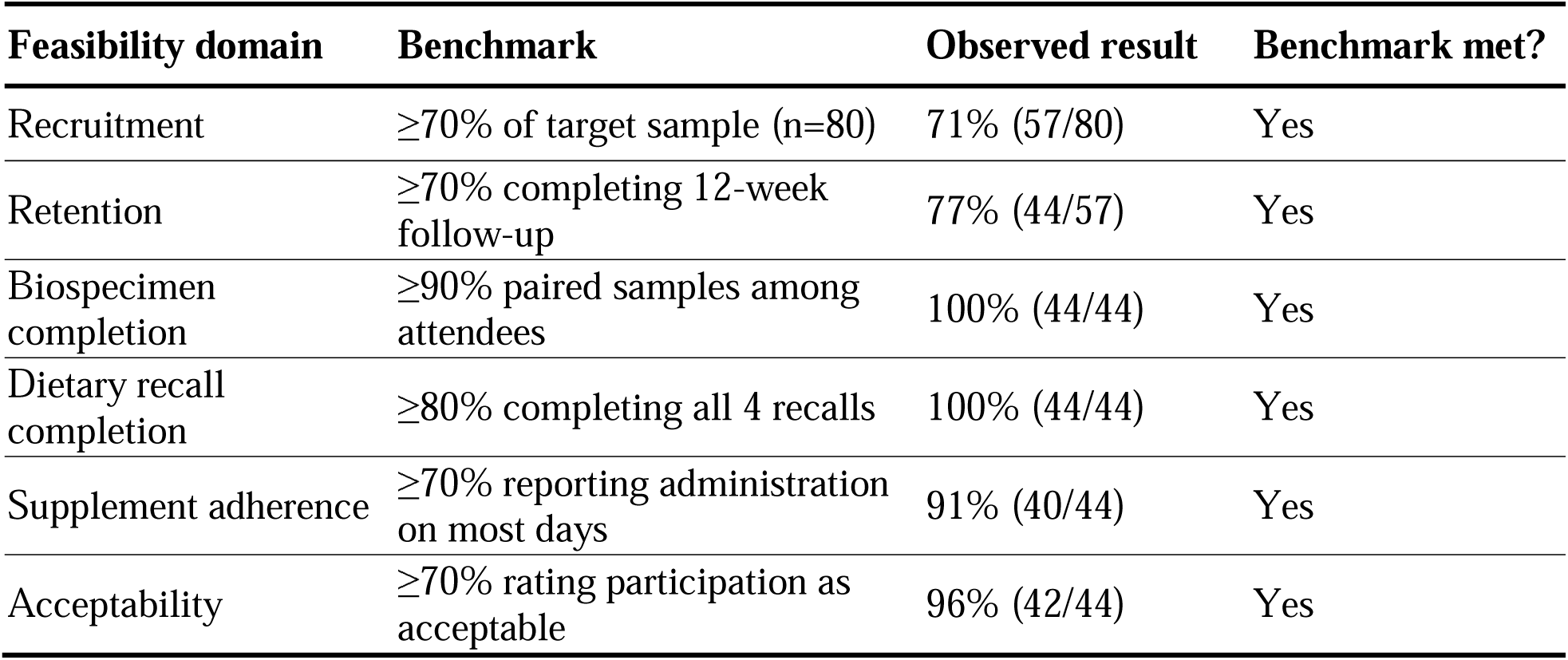
summarizes the feasibility outcomes against their corresponding benchmarks. Biospecimen completion, dietary recall completion, adherence, and acceptability rates reported in Table 3 reflect per-protocol estimates among the 44 participants who completed the 12-week follow-up. When calculated using all 57 enrolled participants as the denominator, these rates are attenuated by the 23% attrition rate; however, no participant who remained in the study failed to complete any planned study procedure, indicating that attrition rather than procedural burden drove the lower intent-to-treat estimates.

| **Feasibility domain** | **Benchmark** | **Observed result** | **Benchmark met?** |
| --- | --- | --- | --- |
| Recruitment | ≥70% of target sample (n=80) | 71% (57/80) | Yes |
| Retention | ≥70% completing 12-week follow-up | 77% (44/57) | Yes |
| Biospecimen completion | ≥90% paired samples among attendees | 100% (44/44) | Yes |
| Dietary recall completion | ≥80% completing all 4 recalls | 100% (44/44) | Yes |
| Supplement adherence | ≥70% reporting administration on most days | 91% (40/44) | Yes |
| Acceptability | ≥70% rating participation as acceptable | 96% (42/44) | Yes |

### Biospecimen Completion

Dried blood spot collection via finger prick was successfully completed at both time points for all participants who attended the baseline and 12-week visits (100% completion among attendees). The procedure was generally well tolerated, though some younger participants required additional time, reassurance, or distraction techniques. The minimally invasive nature of the finger-prick method was an important factor in caregiver willingness to consent to biospecimen collection, and several caregivers noted during follow-up calls that they preferred this approach over venipuncture. Specimen quality was sufficient for laboratory analysis in all collected samples, and the simplified storage and shipping procedures (room temperature storage, batch shipment) were logistically straightforward to implement within the research setting. The biospecimen completion benchmark (≥90%) was met.

### Dietary Recall Completion

All 44 participants who completed the 12-week study finished all four planned 24-hour dietary recalls (two at baseline, two at 12 weeks), yielding 176 total completed recalls among study completers (100% completion). An additional 11 participants who later discontinued completed baseline dietary recalls prior to withdrawal. Despite these high completion rates, scheduling required substantial coordination between nutrition assessment personnel and families. Weekend-day recalls were particularly challenging to schedule, as families were less consistently available for telephone contact on weekends. For younger participants, caregiver involvement in the recall process was essential for accurate reporting, which added time to each recall session. The dietary recall completion benchmark (≥80%) was met.

### Caregiver-Reported Acceptability

Caregiver exit questionnaire responses collected at the 12-week visit indicated high overall acceptability of study participation. The majority of caregivers rated daily supplement administration as acceptable and reported openness to incorporating dietary supplements into their child’s future migraine management. The majority of caregivers rated daily supplement administration as acceptable and reported openness to incorporating dietary supplements into their child’s future migraine management (**Figure 3**). Many caregivers reported observing fewer or less severe migraines during the study period, and some reported improvements in emotional well-being and participation in daily activities. Following study participation, most caregivers reported increased interest in non-pharmacologic migraine management approaches **(Supplemental Tables S1-S4).** The acceptability benchmark (≥70% rating participation as acceptable) was met.

**Figure 3.**
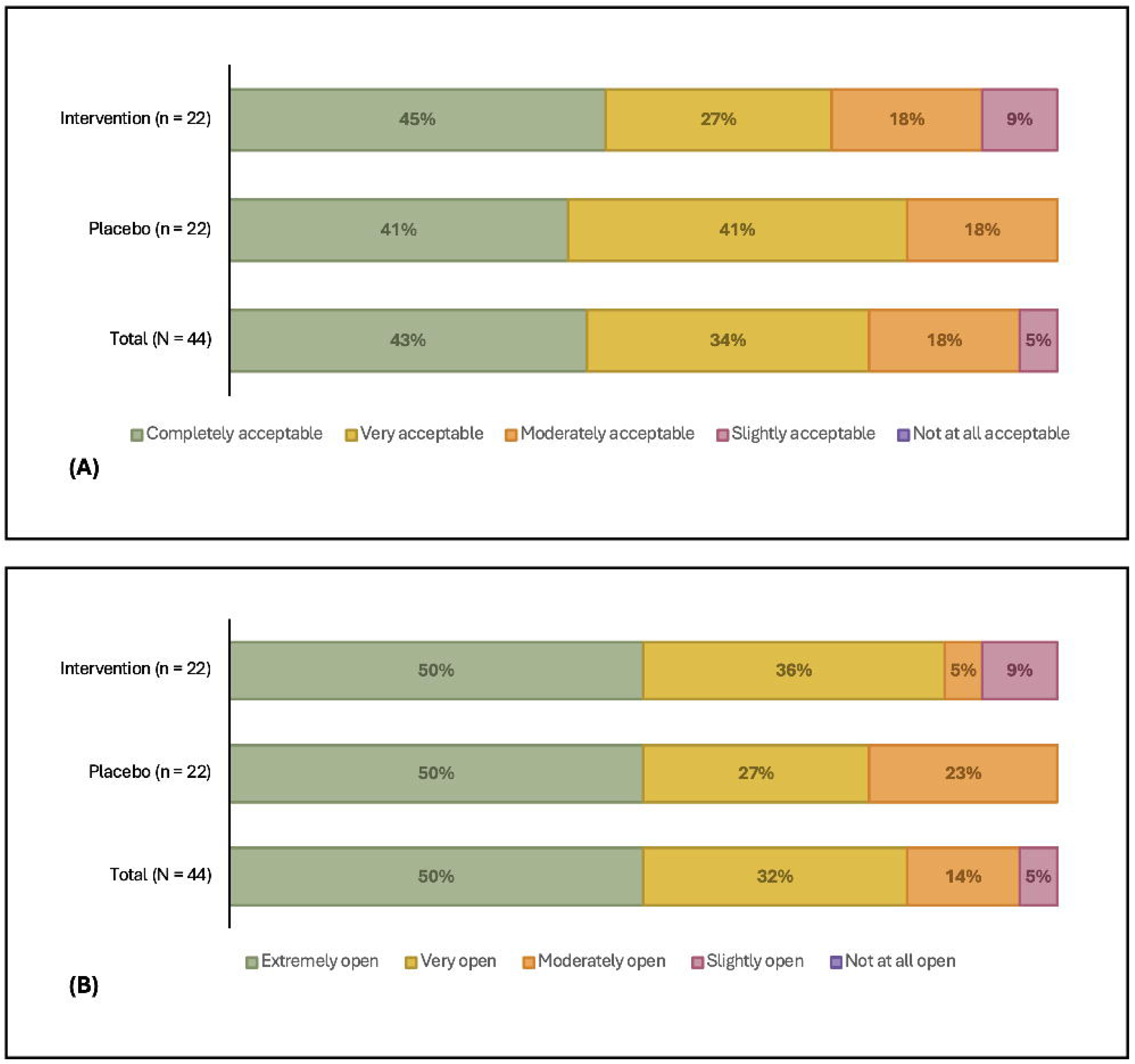
Caregiver-reported acceptability and future openness toward dietary supplementation for pediatric migraine management. (a) Acceptability of daily supplement administration over the 12-week study period; (b) Parent/caregiver openness to including dietary supplements in future migraine management.

### Adherence Monitoring

Caregiver-reported adherence at week 4 and week 8 telephone follow-up calls indicated that the majority of families administered the supplement on most days, meeting the adherence benchmark (≥70% reporting administration on ≥5 of 7 days per week). Most families were able to maintain daily supplement administration throughout the study period, though occasional missed doses were commonly reported. The most frequently cited challenges included forgetting doses during disruptions to daily routine (e.g., travel, weekends, school breaks) and child reluctance related to taste or texture of the liquid formulation. The structured follow-up calls provided an opportunity to troubleshoot administration challenges and reinforce the importance of consistent dosing.

No serious adverse events were reported during the study. Mild tolerability concerns reported during follow-up calls included occasional gastrointestinal symptoms and taste-related reluctance, none of which led to study discontinuation for safety reasons. Two participants discontinued due to supplement palatability, as noted above.

### Protocol Deviations and Practical Lessons

No substantive protocol modifications were required during the study period. During data preparation, a programming error in the electronic data capture system was identified after enrollment was complete: the PedsQL Pediatric Pain Questionnaire had been loaded in place of the Pediatric Quality of Life Inventory (PedsQL) Measurement Model, capturing a construct redundant with the prespecified PROMIS Pediatric Pain Intensity measure. The decision to exclude this outcome was made prior to unblinding and was not motivated by observed outcome data; full details are reported in the trial results manuscript. This experience highlights the importance of independent verification and pilot testing of all instruments within the electronic data capture system prior to enrollment.

Key practical lessons from this feasibility trial include: 1) structured telephone follow-up at regular intervals supports both retention and adherence monitoring with minimal participant burden; 2) dried blood spot collection is well suited to pediatric nutrition research given its minimal invasiveness and simplified logistics; 3) dietary recall scheduling requires dedicated coordination resources, particularly for weekend-day recalls; 4) caregiver engagement is central to all aspects of pediatric supplementation trials, from daily administration to longitudinal follow-up, and study designs should explicitly incorporate caregiver-centered communication and support; and 5) future trials should incorporate prospective daily headache diaries to capture migraine frequency, duration, and severity throughout the intervention period. The present study assessed migraine-related disability retrospectively using the PedMIDAS, which captures functional impairment over the preceding three months but does not provide granular longitudinal data on headache occurrence. Daily electronic headache diaries are considered the preferred method for capturing migraine frequency in clinical trials and would strengthen the ability to detect treatment-related changes in headache patterns, particularly in adequately powered studies where headache days serve as a primary clinical endpoint.

Clinical and biologic efficacy outcomes, including changes in the omega-3 PUFA index, patient-reported outcome measures, and exploratory lipid biomarkers, will be reported in a separate manuscript.

### Summary of Feasibility

All six post-hoc feasibility benchmarks were met.

## Discussion

This study evaluated the feasibility and acceptability of conducting a quasi-randomized, double-blind, placebo-controlled trial of omega-3 PUFA supplementation in children and adolescents with migraine, integrating longitudinal dried blood spot biomarker collection, repeated dietary assessment, validated patient-reported outcome measures, structured remote follow-up, and caregiver-reported acceptability measures. All six post-hoc feasibility benchmarks were met: recruitment reached 71% of the target sample, retention was 77%, caregiver-reported supplement adherence met the ≥70% threshold, biospecimen and dietary recall completion rates were 100% among study completers, and caregiver-reported acceptability was rated moderately acceptable or better by 96% and completely or very acceptable by 77% of study completers. These findings support progression to a future, adequately powered RCT with the design modifications outlined below.

### Methodological Contributions

Several design features of this protocol address recognized challenges in pediatric nutrition intervention research. First, the incorporation of dried blood spot lipid biomarker collection at baseline and 12 weeks provides an objective measure of nutritional target engagement that is independent of self-reported adherence. The ISSFAL has recommended that omega-3 levels be measured at baseline and post-intervention in supplementation trials to confirm biologic response [24]. The minimally invasive finger-prick collection method, requiring only a small volume of capillary blood, was specifically selected to reduce participant burden and improve feasibility in a pediatric population, consistent with evidence supporting dried blood spot methodology in community-based and pediatric research [33,35]. The finger-prick collection method using coated blood cards offers a distinct advantage of preventing oxidation of fatty acids and minimizing blood draws, which can impede recruitment, retention, and repeated biomarker assessment in children and adolescents.

Second, the use of coconut oil as the placebo comparator represents a methodological strength relative to prior omega-3 trials. Olive oil and corn oil placebos have been criticized because they may exert independent immunomodulatory or metabolically active effects that obscure between-group differences [23]. Coconut oil, composed predominantly of medium-chain and saturated fatty acids, does not participate in omega-3 or omega-6 PUFA metabolic pathways and is not expected to modulate eicosanoid or docosanoid signaling relevant to migraine pathophysiology and is therefore considered a more metabolically inert comparator [22]. Although medium-chain triglycerides have been associated with migraine benefit in the context of ketogenic diets through ketone body-mediated metabolic pathways, the dose administered in this study (one teaspoon daily) is substantially below the threshold required to induce meaningful ketogenesis, making clinically relevant metabolic effects of the placebo unlikely [42,43]. This placebo selection may improve the interpretability of both biomarker and clinical outcomes in future trials adopting this framework.

Third, the integration of repeated telephone-administered 24-hour dietary recalls using the Nutrition Data System for Research allowed characterization of background dietary omega-3 intake independent of the study intervention. This is an important methodological strength, as habitual dietary intake can influence both baseline omega-3 status and the magnitude of biomarker response to supplementation [17,18]. Including dietary assessment in future trials may help explain inter-individual variability in biomarker and clinical responses.

Fourth, the structured telephone follow-up procedures at weeks 4 and 8 served multiple functions: monitoring supplement adherence and tolerability, capturing interval changes in medication or medical history, supporting participant retention, and maintaining caregiver engagement. These remote monitoring procedures reduced participant burden by minimizing in-person visits while providing standardized longitudinal data on intervention feasibility. This approach is consistent with recommendations for pediatric trial designs that prioritize family-centered engagement strategies to optimize retention [14,16].

### Recruitment and Multi-Site Coordination

The multi-pathway recruitment strategy, combining data warehouse screening, clinician referral, and clinic-based self-referral across two academic health systems, was designed to maximize identification of eligible participants while accommodating the practical realities of pediatric neurology clinic workflows. Centralizing all study procedures at a single research site while recruiting from two health systems simplified procedural consistency but required careful coordination of communication between research personnel and clinical teams. This model may be applicable to other multi-site pediatric intervention studies where centralized assessment is feasible.

### Caregiver Engagement and Acceptability

The inclusion of a caregiver-reported proxy exit questionnaire represents an important but underutilized component of pediatric intervention trial design. Caregivers play a central role in supplement administration, appointment attendance, and treatment decision-making in pediatric populations, yet caregiver perspectives on intervention acceptability and study burden are infrequently assessed in a structured manner [15]. The caregiver exit questionnaire in this protocol captured caregiver perceptions of supplement acceptability, openness to future dietary supplementation, perceived changes during the study period, and concerns regarding current migraine management options. These data provide important context for interpreting retention and adherence outcomes and for designing future trials that align with family preferences and priorities.

### Limitations and Design Considerations

Several limitations of the study design should be acknowledged. Although alternating allocation maintained numerical balance between study arms, it does not constitute true randomization because treatment assignments are predictable. Future trials should use concealed computer-generated block randomization, which provides both balanced enrollment and protection against allocation bias.

Feasibility objectives were defined prospectively, but formal numerical thresholds were not pre-specified prior to data collection. Benchmarks were established post hoc by reference to published pediatric trial literature. While this approach is transparent and informed by external evidence, pre-specified progression criteria would strengthen the interpretability of feasibility findings in future studies. Notably, a systematic survey of pilot and feasibility studies found that only 4% reported progression criteria at all [44], and a scoping review of 69 pilot/feasibility RCTs found that only 18 reported prespecified progression criteria [45], suggesting that the present approach, while not ideal, reflects common practice in the field. Current consensus guidance acknowledges that not all feasibility parameters require formal benchmarks and that investigators may pre-specify only those most relevant to the planned full-scale trial [46]. Future trials building on this work should establish explicit numerical progression criteria prior to data collection, consistent with current recommendations for pilot and feasibility trial design [19].

Reliance on caregiver report for adherence monitoring without objective measures (e.g., bottle weight, electronic monitoring) is a limitation. Caregiver-reported adherence may overestimate actual supplement consumption, and future larger-scale trials should incorporate objective adherence assessment.

The absence of prospective daily headache diaries is a recognized limitation. Headache frequency assessed through daily diaries provides more granular and reliable data than retrospective recall-based instruments such as the PedMIDAS and is the recommended primary endpoint in migraine clinical trials [9,11,47]. The present study assessed migraine-related disability retrospectively, which does not capture longitudinal headache occurrence during the intervention period. Future trials should incorporate electronic daily headache diaries to capture migraine frequency, duration, and severity prospectively.

Centralizing all study procedures at a single site while recruiting from two health systems facilitated procedural consistency but required participants recruited through Duke to travel to UNC for in-person visits, which may have limited enrollment from that site and contributed to attrition. Future multi-site trials may benefit from distributed study visit capabilities to reduce participant travel burden.

The study population was recruited from academic pediatric neurology clinics, which may limit generalizability to community-based settings or populations with different demographic characteristics. The predominantly White and non-Hispanic composition of the study sample, while reflective of the catchment area, underscores the need for future trials to implement targeted recruitment strategies to ensure adequate representation of diverse populations. Furthermore, there may be a need to assess genetic variability in the response to omega-3 fatty acids, as the host genome controls the metabolism of omega-3 PUFA desaturation and elongation [48].

Finally, the small sample size limits statistical power for detecting between-group differences in patient-reported outcomes and precludes meaningful subgroup analyses. The feasibility and variability data generated by this protocol are intended to inform sample size calculations for future adequately powered trials.

### Future Directions

This study provides a practical, reproducible framework that can be adapted for future pediatric dietary intervention trials. Key design modifications for subsequent studies informed by the present feasibility findings should include: concealed computer-generated randomization with stratification by age and migraine frequency; prospective electronic daily headache diaries as the primary clinical endpoint; longer intervention duration (≥16 weeks) to allow sufficient time for lipid-mediated biologic changes to translate into clinical effects; consideration of higher omega-3 doses informed by predictive modeling of omega-3 index response to supplementation; objective adherence monitoring through bottle weight or electronic tracking; targeted recruitment strategies to improve demographic diversity; and pre-specified feasibility progression criteria with explicit numerical thresholds established prior to data collection, consistent with current recommendations for pilot and feasibility trial design.

The structured telephone follow-up model used in this study effectively supported retention and caregiver engagement with minimal participant burden and should be retained in future trials. Dietary recall scheduling required substantial coordination resources, particularly for weekend-day recalls; future studies may consider whether digital or app-based dietary assessment tools could reduce scheduling burden while maintaining data quality. Caregiver engagement was central to all aspects of this trial, from daily supplement administration to longitudinal follow-up, and future study designs should explicitly incorporate caregiver-centered communication and support strategies.

Distributed study visit capabilities across recruitment sites may also improve enrollment in future multi-site trials, as centralizing all procedures at a single site, while supporting procedural consistency, required some participants to travel to a separate institution for in-person visits.

## Conclusions

This study demonstrates that a quasi-randomized, double-blind, placebo-controlled trial of omega-3 PUFA supplementation in youth with migraine integrating longitudinal dried blood spot biomarker collection, repeated dietary assessment, validated patient-reported outcome measures, structured remote follow-up, and caregiver-reported acceptability measures is feasible and acceptable to families. All six post-hoc feasibility benchmarks were met: recruitment reached 71% of the target sample, retention was 77%, caregiver-reported supplement adherence met the ≥70% threshold, biospecimen and dietary recall completion rates were 100% among study completers, and caregiver-reported acceptability was rated moderate or better among 96% of study completers. Based on these findings, a future adequately powered RCT is warranted and is currently being planned, incorporating concealed randomization, pre-specified progression criteria, prospective daily headache diaries, longer intervention duration, objective adherence monitoring, and targeted recruitment strategies to improve demographic diversity. The methodological framework and practical lessons described herein may inform the design and implementation of future larger-scale pediatric dietary intervention trials evaluating omega-3 supplementation and other nutrition-based interventions in youth with migraine.

## Data Availability

All data produced in the present study are available upon reasonable request to the authors.

## Abbreviations

AA: arachidonic acid
ANCOVA: Analysis of Covariance
CDART: Carolina Data Acquisition and Reporting Tool
CONSORT: Consolidated Standards of Reporting Trials
DHA: docosahexaenoic acid
EPA: eicosapentaenoic acid
HIPAA: Health Insurance Portability and Accountability Act
ICHD-3: International Classification of Headache Disorders, 3rd edition
IRB: Institutional Review Board
ISSFAL: International Society for the Study of Fatty Acids and Lipids
NC TraCS: NC Translational and Clinical Sciences Institute
NCATS: National Center for Advancing Translational Sciences
NDSR: Nutrition Data System for Research
PedMIDAS: Pediatric Migraine Disability Assessment
PedsQL: Pediatric Quality of Life Inventory
PROMIS: Patient-Reported Outcomes Measurement Information System
PUFA: polyunsaturated fatty acids
RCADS: Revised Children’s Anxiety and Depression Scale
RCT: randomized controlled trial
UNC: University of North Carolina

## Declarations

### Ethics Approval and Consent to Participate

The study was conducted in accordance with the Declaration of Helsinki and approved by the Institutional Review Board of University of North Carolina (IRB#24-2715, 26 February 2025) for studies involving humans. Informed consent was obtained from all subjects involved in the study. Caregivers or legal guardians of participants were re-consented or notified in the event that new information pertinent to the study became available, previously unrecognized risks were identified, significant modifications to the study protocol were implemented, or deficiencies in the initial consent process were determined.

### Consent for Publication

This is included as part of the participant consent and assent forms.

### Availability of Data and Materials

The original contributions presented in this study are included in the article/supplementary material. Further inquiries can be directed to the corresponding author.

### Authors’ Contributions

Conceptualization, C.M.S. and A.S.; methodology, C.M.S., D.Z., K.F., R.S..; formal analysis, A.S., K.D.; investigation, S.G., K.W., S.T.; resources, C.M.S..; data curation, S.G., K.W., S.T., J.B.; writing—original draft preparation, J.B., C.M.S..; writing— review and editing, S.G., A.S., R.S., K.D., K.F., D.Z., K.W., S.T.; visualization, A.S., K.D., J.B.; supervision, C.M.S.; funding acquisition, C.M.S. All authors have read and agreed to the published version of the manuscript.

## Acknowledgements

We acknowledge Nordic Naturals for generously providing the study intervention used in this trial. We also thank the UNC Nutrition and Obesity Research Center Clinical and Community Human Assessment and Intervention Core: Behavioral Assessment for assistance with dietary recalls. We also acknowledge Patience Ben-Israel, Sara La Lone, Sydney Danze, Sajan Singh, Thomas Southern, and Joseph Iskander for their assistance with participant recruitment. Finally, we acknowledge the assistance of the NC Translational and Clinical Sciences (NC TraCS) Institute, which is supported by the National Center for Advancing Translational Sciences (NCATS), National Institutes of Health, through Grant Award Number UM1TR004406, for assistance with participant identification through the Carolina Data Warehouse for Health.

## Supplemental Materials

**Supplemental File 1.** Completed CONSORT checklist.

**Supplemental Table S1.** Caregiver-Reported Access to Non-Pharmacologic Migraine Treatments Prior to Study Participation

**Supplemental Table S2.** Change in Caregiver Interest in Non-Pharmacologic Migraine Treatments Following Study Participation

**Supplemental Table S3.** Caregiver-Reported Benefits Observed During Study Participation

**Supplemental Table S4.** Caregiver Concerns Regarding Current Pediatric Migraine Treatment Options

