## Supplemental Tables S1-S4 for "Omega-3 Supplementation with Lipid Biomarker Assessment in Youth with Migraine: A Quasi-Randomized Controlled Feasibility Trial"

**Table S1. Caregiver-Reported Access to Non-Pharmacologic Migraine Treatments Prior to Study Participation**

| | Very easy | Somewhat easy | Neither easy nor difficult | Somewhat difficult | Very difficult |
|---|---|---|---|---|---|
| **Total (N = 44)** | 15 (34.1%) | 16 (36.4%) | 7 (15.9%) | 5 (11.4%) | 1 (2.3%) |
| **Placebo (n = 22)** | 8 (36.4%) | 7 (31.8%) | 3 (13.6%) | 4 (18.2%) | 0 (0.0%) |
| **Intervention (n = 22)** | 7 (31.8%) | 9 (40.9%) | 4 (18.2%) | 1 (4.5%) | 1 (4.5%) |

Data are presented as the number and percentage of respondents in each category for the total sample (N = 44) and by group (Placebo, n = 22; Intervention, n = 22).

**Table S2. Change in Caregiver Interest in Non-Pharmacologic Migraine Treatments Following Study Participation**

| | Much more interested | Slightly more interested | No change | Slightly less interested | Much less interested |
|---|---|---|---|---|---|
| **Total (N = 44)** | 14 (31.8%) | 15 (34.1%) | 13 (29.5%) | 2 (4.5%) | 0 (0.0%) |
| **Placebo (n = 22)** | 9 (40.9%) | 7 (31.8%) | 5 (22.7%) | 1 (4.5%) | 0 (0.0%) |
| **Intervention (n = 22)** | 5 (22.7%) | 8 (36.4%) | 8 (36.4%) | 1 (4.5%) | 0 (0.0%) |

Data are presented as the number and percentage of respondents in each category for the total sample (N = 44) and by group (Placebo, n = 22; Intervention, n = 22).

**Table S3. Caregiver-Reported Benefits Observed During Study Participation**

| | Fewer or less severe migraines | Improved emotional well-being | Better ability to participate in daily activities | Improved quality of life for our family | Other | No change |
|---|---|---|---|---|---|---|
| **Total (N = 44)** | 27 (61.4%) | 13 (29.5%) | 11 (25.0%) | 8 (18.2%) | 5 (11.4%) | 7 (15.9%) |
| **Placebo (n = 22)** | 13 (59.1%) | 5 (22.7%) | 6 (27.3%) | 3 (13.6%) | 2 (9.1%) | 3 (13.6%) |
| **Intervention (n = 22)** | 14 (63.6%) | 8 (36.4%) | 5 (22.7%) | 5 (22.7%) | 3 (13.6%) | 4 (18.2%) |

Data are presented as the number and percentage of respondents in each category for the total sample (N = 44) and by group (Placebo, n = 22; Intervention, n = 22).
Respondents could select multiple options; therefore, percentages exceed 100%.

**Table S4. Caregiver Concerns Regarding Current Pediatric Migraine Treatment Options**

| | No major concerns | We prefer to avoid medications if possible | Hard for my child to tolerate | Difficult to access or afford | Not effective enough | Too many side effects | Other |
|---|---|---|---|---|---|---|---|
| **Total (N = 44)** | 25 (56.8%) | 4 (9.1%) | 2 (4.5%) | 3 (6.8%) | 11 (25.0%) | 3 (6.8%) | 5 (11.4%) |
| **Placebo (n = 22)** | 12 (54.5%) | 1 (4.5%) | 2 (9.1%) | 1 (4.5%) | 7 (31.8%) | 2 (9.1%) | 3 (13.6%) |
| **Intervention (n = 22)** | 13 (59.1%) | 3 (13.6%) | 0 (0.0%) | 2 (9.1%) | 4 (18.2%) | 1 (4.5%) | 2 (9.1%) |

Data are presented as the number and percentage of respondents in each category for the total sample (N = 44) and by group (Placebo, n = 22; Intervention, n = 22).
Respondents could select multiple options; therefore, percentages exceed 100%.
